# COVID-19 Spreading Dynamics in an Age-Structured Population with Selective Relaxation of Restrictions for Vaccinated Individuals : a Mathematical Modeling Study

**DOI:** 10.1101/2021.02.22.21252241

**Authors:** B Shayak, Mohit M Sharma, Anand K Mishra

**Affiliations:** Mechanical and Aerospace Engineering, Cornell University, Ithaca – 14853, New York State, USA; Population Health Sciences, Weill Cornell Medicine, 1300 York Avenue, New York City – 10065, New York State, USA

**Keywords:** Immunity passport, Vaccine efficacy, Viral strain, Delay differential, equation, Age-structuring

## Abstract

**Background:** COVID-19 vaccination of healthcare and other essential workers is underway in many countries while immunization of the general public is expected to begin in the next several weeks. We consider the question of whether people who have received the vaccine can be selectively and immediately permitted to return to normal activities.

**Methods:** We use a delay differential equation model to calculate the effects of vaccinee “immunity passports” on the epidemic spreading trajectories. The model incorporates age-structuring to account for children who are ineligible for vaccination, and senior citizens who are especially vulnerable to the disease. We consider consensus strains of virus as well as high-transmissibility variants such as B1.1.7 and B1.351 in our analysis.

**Results:** We find that with high vaccine efficacy of 80 percent or greater, unrestricted vaccinee—vaccinee interactions do not derail the epidemic from a path towards elimination. Vaccinee—non-vaccinee interactions should however be treated with far more caution. At current vaccine administration rates, it may be the better part of a year before COVID-19 transmission is significantly reduced or ceased. With lower vaccine efficacy of approximately 60 percent, restrictions for vaccinees may need to remain in place until the elimination of the disease is achieved. In all cases, the death tolls can be reduced by vaccinating the vulnerable population first.

**Conclusions:** Designing high-efficacy vaccines with easily scalable manufacturing and distribution capacity should remain on the priority list in academic as well as industrial circles. Performance of all vaccines should continue to be monitored in real time during vaccination drives with a view to analysing socio-demographic determinants of efficacy, if any, and optimizing distribution accordingly. A speedy and efficacious vaccination drive augmented with selective relaxations for vaccinees will provide the smoothest path out of the pandemic with the least additional caseloads, death tolls and socio-economic cost.

## Introduction

Over the past couple of months, four COVID-19 vaccine candidates – the ones developed by Pfizer/ BioNTech, Moderna, Oxford/ Astra Zeneca (also Oxford/ Serum Institute of India) and ICMR/ Bharat Biotech – have received emergency use authorization following rigorous trial procedures. These are being used in vaccination drives all over the world; currently, healthcare workers and essential workers or vulnerable populations are the beneficiaries, with vaccination of the general public being on the to-do list. The first two vaccine candidates (mRNA vaccines)^1,2^ have reported trial efficacies of almost 95 percent, the third candidate (a vector vaccine)^3^ has reported 60-90 percent (dosage-dependent), while the fourth (an adjuvated inactivated vaccine)^4,5^ has reported encouraging immunogenicity results in the early trials; participant enrolment for the phase 3 trial has been completed but the trial itself has not. In August 2020, Russia approved Sputnik 5 (a vector vaccine) bypassing some of the trial protocols; a recent study^6^ finds its efficacy to be 90 percent. Very recently, Johnson and Johnson (a vector vaccine)^7^ has reported 57-72 percent efficacy (location-dependent) while Novavax (an adjuvated subunit vaccine)^8^ has reported 60 (South Africa) to 90 (UK) percent efficacy; both vaccines are currently undergoing the approval process.

Mathematical modeling studies of COVID-19 dynamics post-vaccination started emerging as soon as the first vaccines were approved. Swan, Goyal, Bracis et. al.^9^ have performed a detailed analysis of the roles played by different vaccine efficacy metrics. Several studies^10–13^ find that vaccinating high-contact people first will have the greatest beneficial effect on the spread of the disease. Foy, Wahl, Mehta et. al.^14^ find that priority vaccination of elderly and vulnerable people is optimal for minimizing deaths. They also find that continuing with social restrictions such as six-foot separation and mask regulations during the vaccination drive will best mitigate the disease spread. This conclusion has been obtained in many other analyses as well^12,15–18^. Some studies however^19,20^ project a more optimistic outcome, permitting a gradual relaxation of non-pharmaceutical interventions (NPI) starting from the fourth month of the vaccination drive.

From the general public’s perspective, continued social restrictions for vaccinees may appear inconvenient. Those who have got the vaccine would at least hope to socialize freely with others who have been vaccinated as well. From an economic perspective, continuing NPI during the vaccination drive will amount to prolonged strain on businesses and on the government’s fiscal resources – any relaxation or exemption will act as a lifeline. Our quest here is to find such an exemption – specifically, we ask whether social restrictions can be immediately and preferentially relaxed for those individuals who have been vaccinated. Hereafter, we refer to this strategy as **“selective relaxation”**. To clarify, we treat a person as vaccinated only after s/he has received the second dose of a two-dose regimen and cleared the subsequent immunogenicity period.

With ideal vaccines, the success of the selective relaxation strategy would have been a given. However, the actual COVID-19 vaccines are not 100 percent efficacious, which raises the issue of whether unrestricted (or at least significantly expanded) social activity and mobility on the part of vaccinees may drive the epidemic out of control. In the remainder of this Article, we use mathematical modeling to address this question.

## Methods

We use a compartmental or lumped parameter delay differential equation model developed by our group^21^. We have selected this model because all parameters here are directly related to the disease or to control measures, and because it can be easily extended to accommodate vaccination.

We present here only the outline of the model, with the full equations and derivation being given in §1 of the Supplementary Data. Time is measured in days. We incorporate age-structuring, defining three groups of people : Group 1 or “minors” aged 0-19, Group 2 or “middlers” aged 20-64 and Group 3 or “seniors” aged 65 and above (the labels “minors”, “middlers” and “seniors” are just for easy identification of the groups). We believe that this is the simplest structure which can account for the facts that (*a*) under-18 (entire Group 1 in our model) are currently ineligible to be vaccinated, (*b*) interaction rates among different age groups are highly disparate, and (*c*) older people are disproportionately more vulnerable to the disease. We have taken the population fractions from USA data^22^.

The entry *q*_*ij*_ (*i,j*=1,2,3) of the contact matrix **Q** denotes the (average) number of people belong to Group *j* with whom one person belonging to Group *i* interacts in one day. We have calculated **Q** based on Mossong, Hens, Jit et. al.^23^; it is

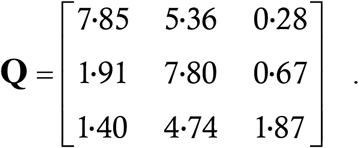

The spreading matrix **M** (*m*_*ij*_ denotes the number of Group *j* targets to whom one at large Group *i* case transmits the infection in one day) is obtained via multiplication of **Q** by a constant *P*_0_. This number denotes the probability that an interaction between a case and a susceptible target actually results in a transmission. Given **Q**, the reproduction number *R*_0_ is proportional to *P*_0_ or to **M**. To represent normal life we use a matrix **M**^*h*^ (‘*h*’ denotes high) adjusted to yield an *R*_0_ of our choice; to represent social restrictions we multiply **M**^*h*^ by the derate *δ* (scalar) to form the matrix **M**^*l*^ (‘*l*’ denotes low). The derate can represent a lower interaction rate (lockdown), lower transmission probability (masking), or (more practically) a combination of the two. We have described this process in detail in §1 of the Supplementary Data.

The major parameters of interest are as follows.

- **Vaccine efficacy *η* :** We define this as the probability that the vaccine works. We assume that when the vaccine does work, it confers full sterilizing immunity – while the vaccines’ efficacy against symptomatic disease is well-known, a recent very important study^24^ has demonstrated the Pfizer vaccine to be effective against asymptomatic infection as well. We also assume that the vaccine confers zero transmissibility-reducing immunity when it does not work.
- **Viral strain :** This informs the choice of the matrix **M**^*h*^; we adjust it so that in the absence of restrictions, the disease has a starting reproduction number *R*_0_ of 3·0, 2·0 or 5·0, corresponding to consensus^25–28^, low-transmissibility and high-transmissibility or rogue strains (such as B1.1.7 and B1.351)^29,30^ of the virus respectively.
- **Interaction mode :** Minors are not eligible for vaccination while middlers and seniors are. As time goes on, more and more of the latter will get the shots and become eligible for relaxations. We consider two modes in which the relaxation takes place. In **Mode 1**, vaccinees resort to the high spreading rates 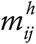 only with other vaccinees and adhere to the low rates 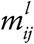 in all other situations, including interactions with minors. The latter being unvaccinated also remain subject to restrictions. This corresponds to an ideal mode of operation where vaccinees relax precautions such as six-foot separation and masking only in the presence of other vaccinees. Interaction Mode 1 can be realized by making proof of vaccination mandatory for admission to unmasked and un-distanced classrooms, conference halls, concerts, sporting events, cinema halls, bars, restaurants and similar venues, while keeping interventions active at full strength everywhere else. **Mode 2** represents a greater degree of relaxation where vaccinees take recourse to the high rates 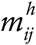 with everyone, independent of their vaccination status. Minors are also permitted the high spreading rates when interacting with vaccinated middlers and seniors. Given the level of intervention fatigue already in society, an attempt to maintain Mode 1 might well drift closer to Mode 2 in reality. For both interaction modes, we choose the initial derate *δ*_0_ such that *R*_0_ equals unity; we then increase *δ* in discrete steps of *δ*_0_/20 every 50 days.

We solve the model using numerical integration in the software Matlab. The method is 2^nd^ order Runge Kutta with a time step of 1/1000 day. The spatial domain of solution is a Notional City of total population 3,00,000. We must seed the model with initial case and vaccination histories defined in the time interval *t* belongs to [0, 7] (days); we choose these functions as 75*t* cases among minors, 175*t* cases among unvaccinated middlers and 50*t* cases among unvaccinated seniors, with zero cases in the vaccinated groups; we take the vaccinee counts as constant function 100 in both eligible groups. We terminate the simulation run and declare the epidemic to be over if the region has less than one active case for fourteen consecutive days. Details of the initial and terminal conditions are given in the Supplementary Data.

To calculate the death tolls, we have used data from O’Driscoll, Dos Santos, Wang et. al.^31^ together with our population structure to obtain the mortality rates of 0·001996 percent, 0·1376 percent and 3·335 percent for minors, middlers and seniors respectively. We are aware that every individual death is a tragedy, and do not seek to undermine this fact while presenting mortality counts as statistics, along with everything else. We assume (currently without quantitative basis) that mortality rates are reduced by a factor of 10 for vaccinees.

## Results

We start by running simulations for selective relaxation with Interaction Mode 1. Let vaccines be distributed at the constant rate 600/day, corresponding to vaccination of the entire population in 500 days. Let 90 percent of the available vaccines be allotted to seniors until they have been vaccinated completely, after which the entire stock is given to middlers. We consider three different vaccine efficacies of 60, 80 and 90 percent and the three viral strains mentioned in the Methods Section. In each case, we use the following metrics to calibrate the region’s infection control performance : (*a*) the time *T* to the end of the outbreak, (*b*) the total number *X* of cases at the end of the outbreak, (*c*) the total number *D* of deaths at the end of the outbreak and (*d*) the vaccination fault ratio *f*, defined as the maximum ratio of vaccinated cases to total vaccinees at any point in the outbreak and reported as a percentage. We present the results in Table 1.

**Table 1 :**
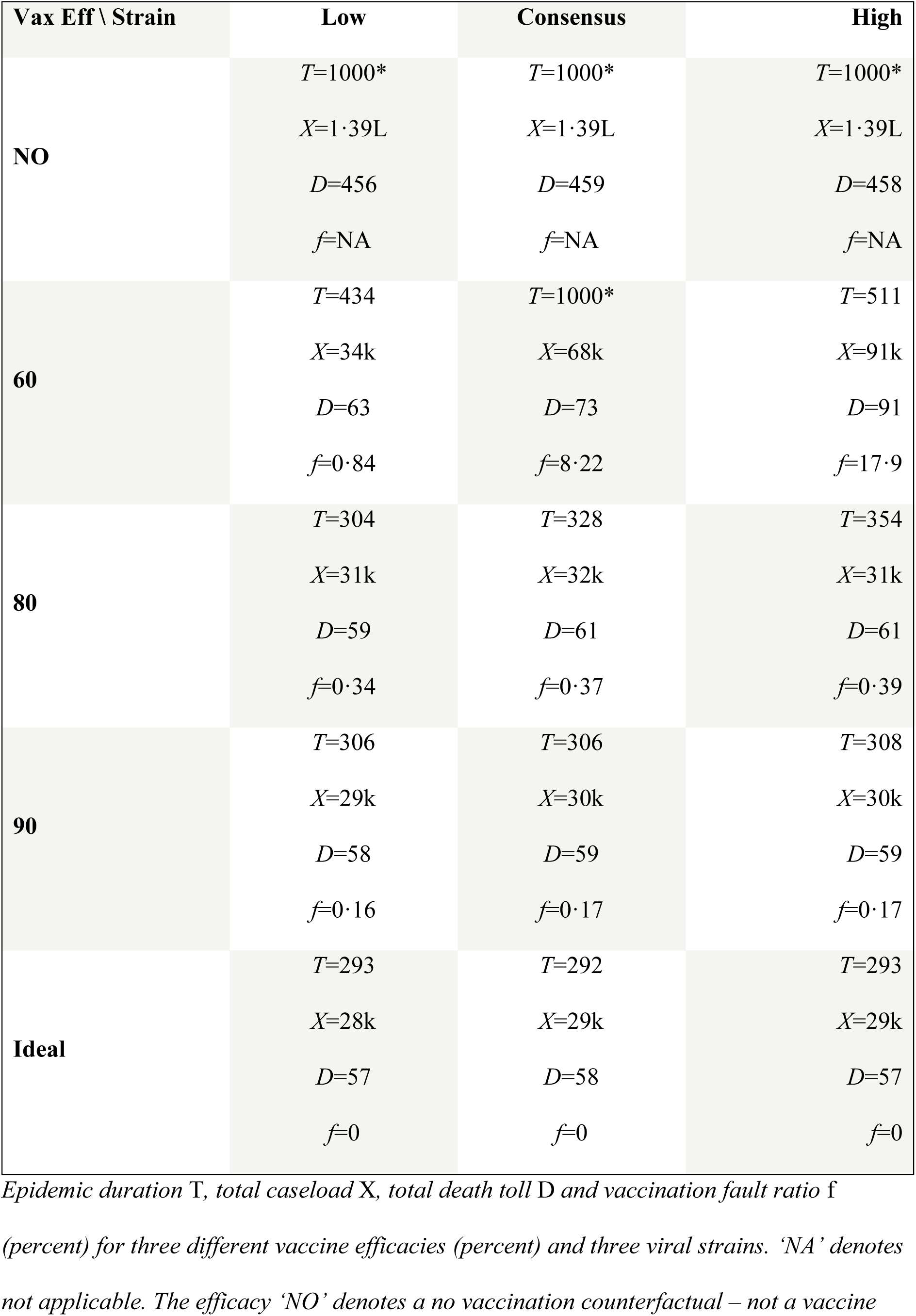
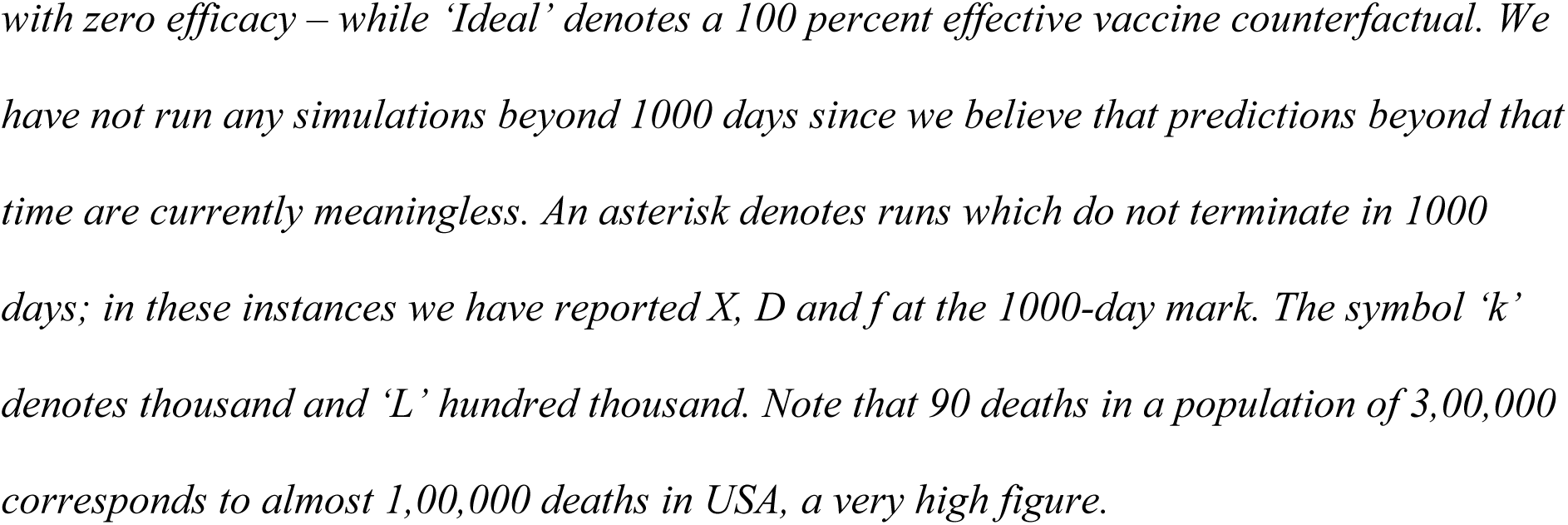
Interaction Mode 1.

| **Vax Eff \ Strain** | **Low** | **Consensus** | **High** |
| --- | --- | --- | --- |
| **NO** | $T=1000^*$ | $T=1000^*$ | $T=1000^*$ |
| | $X=1.39L$ | $X=1.39L$ | $X=1.39L$ |
| | $D=456$ | $D=459$ | $D=458$ |
| | $f=NA$ | $f=NA$ | $f=NA$ |
| **60** | $T=434$ | $T=1000^*$ | $T=511$ |
| | $X=34k$ | $X=68k$ | $X=91k$ |
| | $D=63$ | $D=73$ | $D=91$ |
| | $f=0.84$ | $f=8.22$ | $f=17.9$ |
| **80** | $T=304$ | $T=328$ | $T=354$ |
| | $X=31k$ | $X=32k$ | $X=31k$ |
| | $D=59$ | $D=61$ | $D=61$ |
| | $f=0.34$ | $f=0.37$ | $f=0.39$ |
| **90** | $T=306$ | $T=306$ | $T=308$ |
| | $X=29k$ | $X=30k$ | $X=30k$ |
| | $D=58$ | $D=59$ | $D=59$ |
| | $f=0.16$ | $f=0.17$ | $f=0.17$ |
| **Ideal** | $T=293$ | $T=292$ | $T=293$ |
| | $X=28k$ | $X=29k$ | $X=29k$ |
| | $D=57$ | $D=58$ | $D=57$ |
| | $f=0$ | $f=0$ | $f=0$ |

The case trajectories in the simulations of Table 1 can show either a unimodal or a bimodal profile. We display representative time traces of both kinds in Figure 1.

**Figure 1 :**
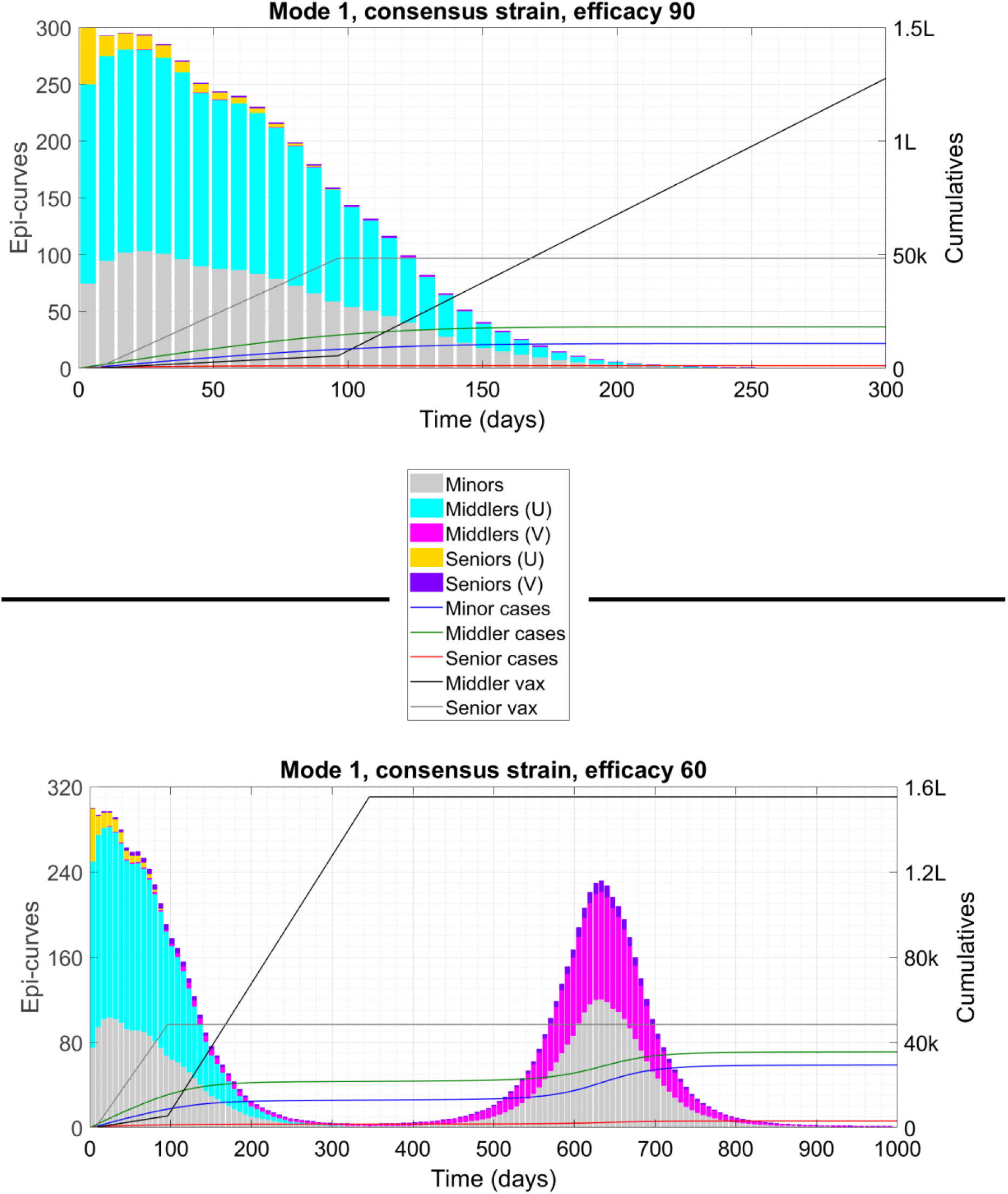
Time traces of two situations from Table 1. The top panel shows a unimodal solution (one peak at t=0) while the bottom panel shows bimodal solution (two peaks). In each plot we display cumulative counters such as caseload and vaccinee count as lines associated with the right-hand y-axis and the weekly case counts or epidemiological curve (epi-curve) as bars associated with the left-hand y-axis. We have scaled down the weekly counts by a factor of 7 so that the envelope of the bars coincides with the time derivative of the cumulative cases. The symbol ‘U’ denotes unvaccinated, ‘V’ vaccinated, ‘k’ thousand and ‘L’ hundred thousand.

We now repeat the exercise of Table 1 but with Interaction Mode 2.

In Tables 1 and 2, a 60 percent effective vaccine seems to be of limited utility while a 90 percent effective vaccine performs well almost always. This raises the question of what constitutes a minimum acceptable efficacy. To analyse this, we consider two different combinations of viral strain and interaction mode, vary the efficacy continuously between 50 and 100 percent, and plot the resulting duration and caseload in the two panels of Figure 2.

**Table 2 :**
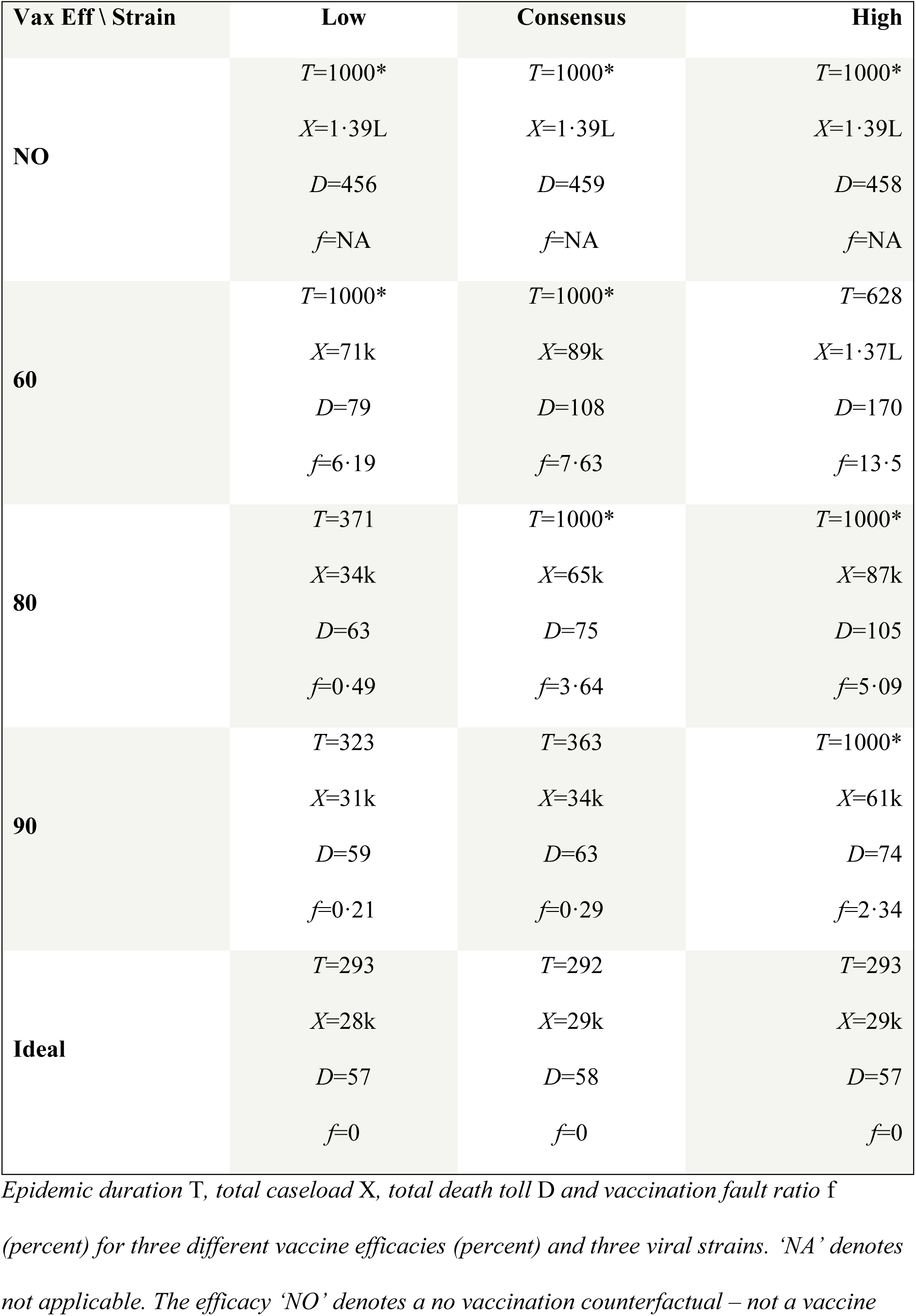
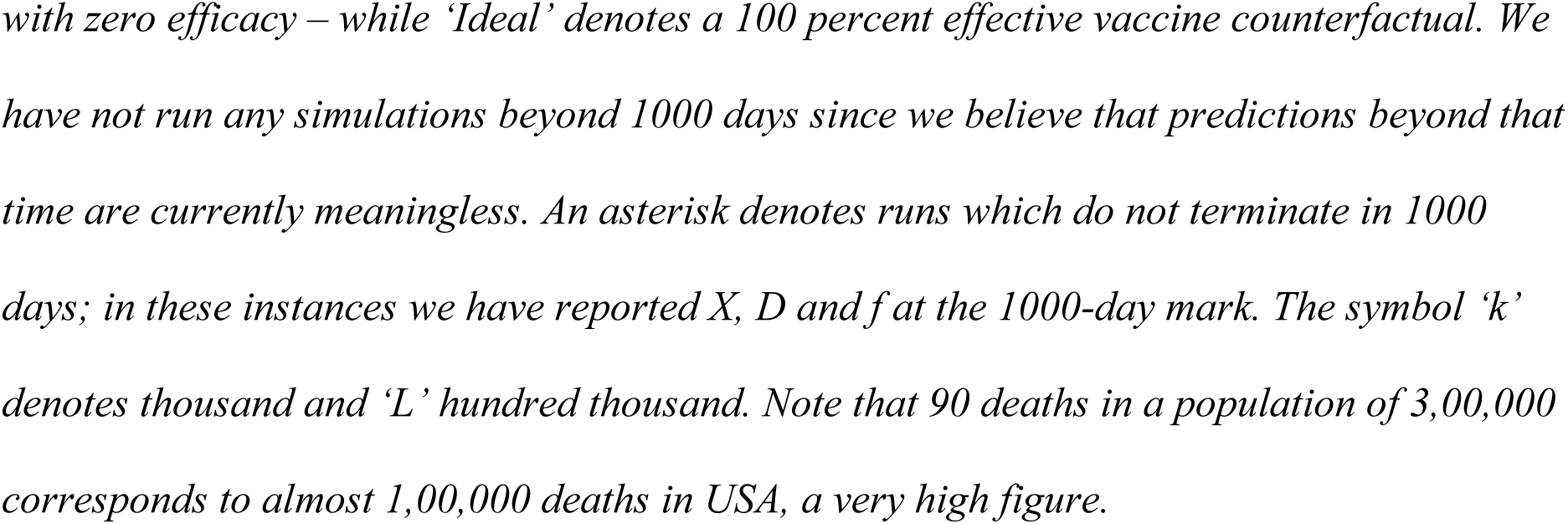
Interaction Mode 2.

| **Vax Eff \ Strain** | **Low** | **Consensus** | **High** |
| --- | --- | --- | --- |
| **NO** | $T=1000^*$ | $T=1000^*$ | $T=1000^*$ |
| | $X=1.39L$ | $X=1.39L$ | $X=1.39L$ |
| | $D=456$ | $D=459$ | $D=458$ |
| | $f=NA$ | $f=NA$ | $f=NA$ |
| **60** | $T=1000^*$ | $T=1000^*$ | $T=628$ |
| | $X=71k$ | $X=89k$ | $X=1.37L$ |
| | $D=79$ | $D=108$ | $D=170$ |
| | $f=6.19$ | $f=7.63$ | $f=13.5$ |
| **80** | $T=371$ | $T=1000^*$ | $T=1000^*$ |
| | $X=34k$ | $X=65k$ | $X=87k$ |
| | $D=63$ | $D=75$ | $D=105$ |
| | $f=0.49$ | $f=3.64$ | $f=5.09$ |
| **90** | $T=323$ | $T=363$ | $T=1000^*$ |
| | $X=31k$ | $X=34k$ | $X=61k$ |
| | $D=59$ | $D=63$ | $D=74$ |
| | $f=0.21$ | $f=0.29$ | $f=2.34$ |
| **Ideal** | $T=293$ | $T=292$ | $T=293$ |
| | $X=28k$ | $X=29k$ | $X=29k$ |
| | $D=57$ | $D=58$ | $D=57$ |
| | $f=0$ | $f=0$ | $f=0$ |

**Figure 2 :**
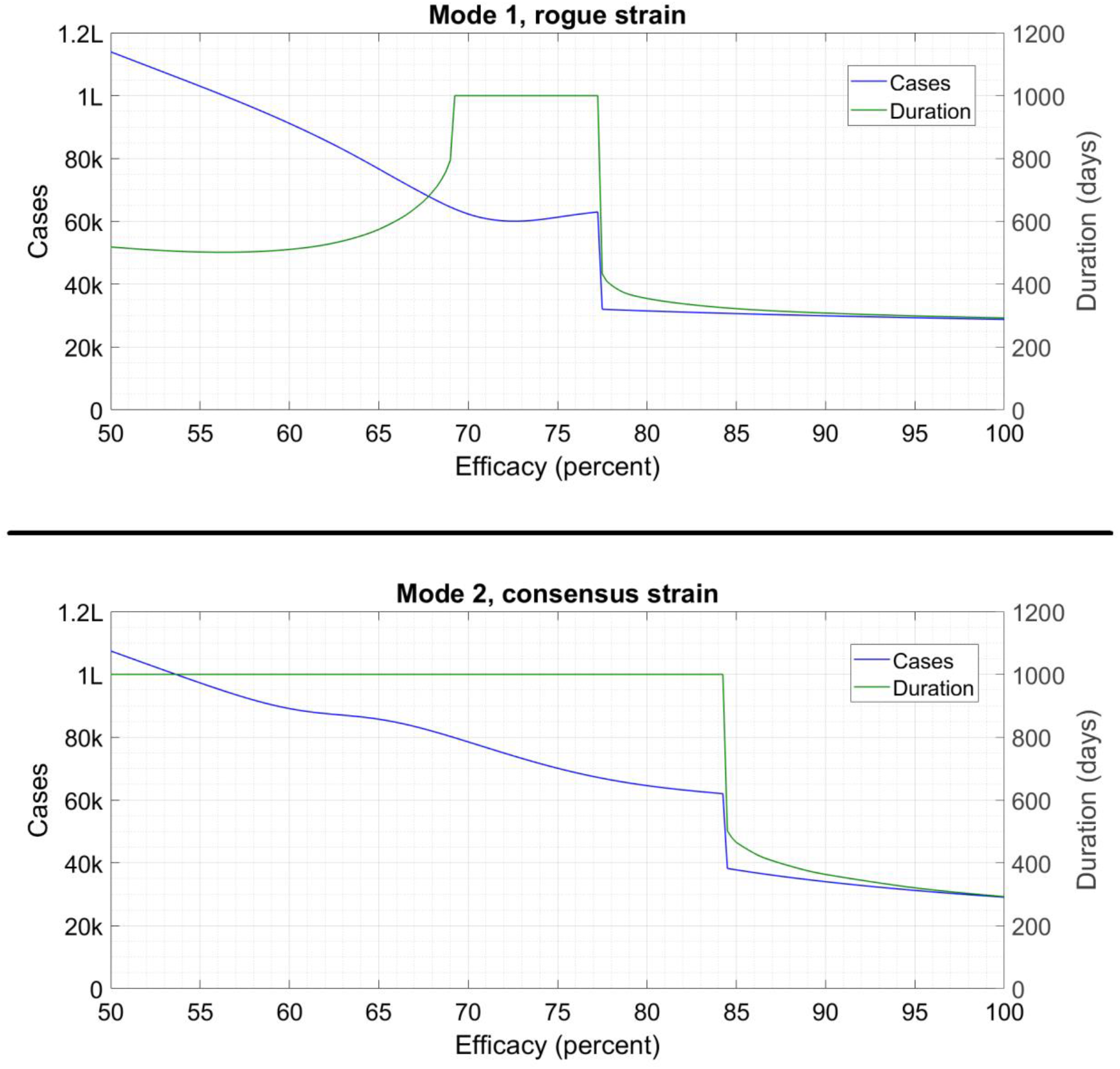
Caseload (left-hand y-axis) and duration (right-hand y-axis) as a function of vaccine efficacy for two combinations of interaction mode and viral strain. The durations have flat regions at 1000 days since we terminate all simulations at this time; in actuality the runs in these regions go on for even longer. The jump in the curves corresponds to a transition from a unimodal to a bimodal solution profile. The symbol ‘k’ denotes thousand and ‘L’ hundred thousand.

For a given vaccine efficacy, one way of achieving lower caseloads and accelerating the end of the pandemic is by increasing the vaccination rate. Considering one combination of interaction mode, viral strain and efficacy, we plot the duration and caseload as a function of vaccination rate in Figure 3.

**Figure 3 :**
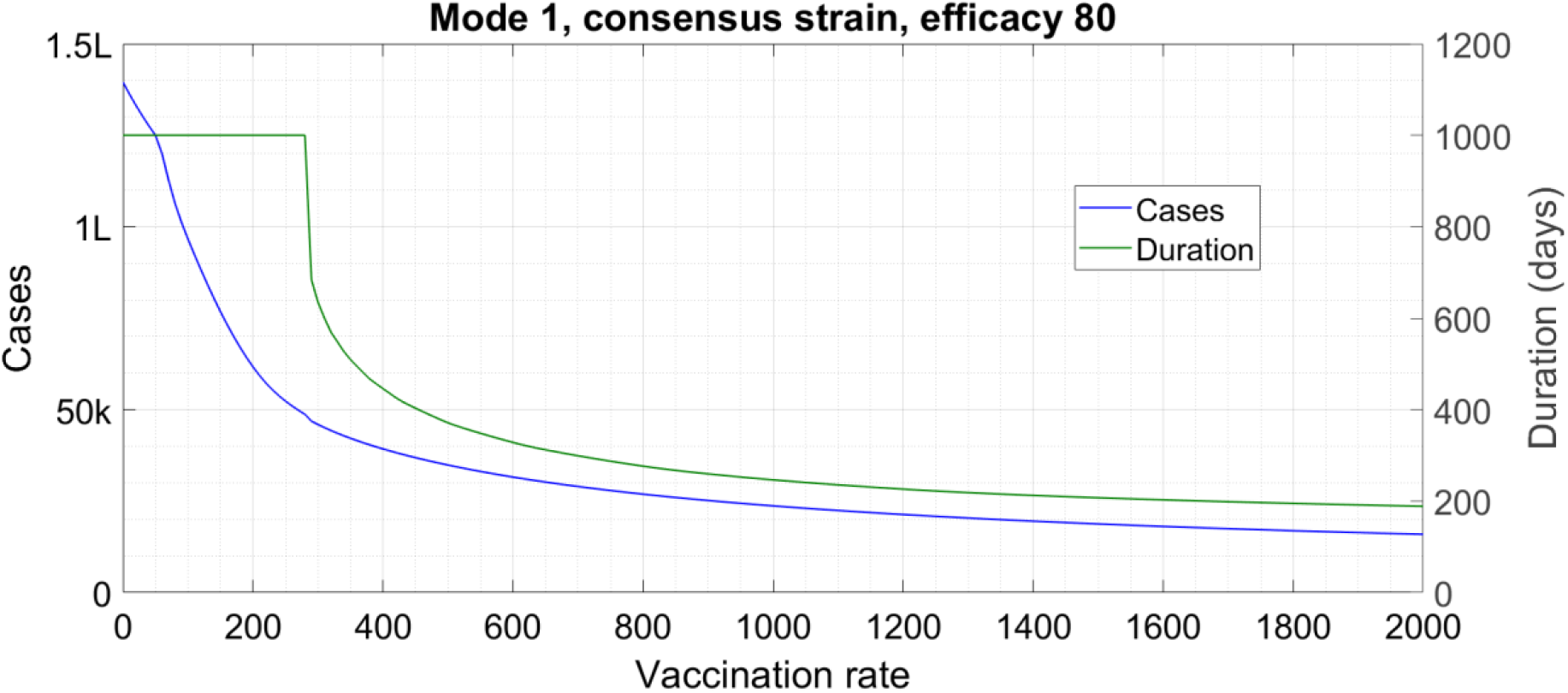
Caseload (left-hand y-axis) and duration (right-hand y-axis) as a function of vaccination rate for a given combination of the other parameters. The duration has a flat region at 1000 days since we terminate all simulations at this time; in actuality the runs in this region go on for even longer. The knee in the curve corresponds to a transition from a unimodal to a bimodal solution profile. The symbol ‘k’ denotes thousand and ‘L’ hundred thousand.

Finally, another way of improving the outcome is by reducing or forgoing the stepwise increments in *δ* every 50 days. Instead of the 5 percent increment, if we go with constant derate *δ*_0_ then the duration as well as caseload can reduce significantly. Considering the consensus viral strain, with Interaction Mode 2, the cutoff vaccine efficacy at which the unimodal solution breaks down is approximately 74 percent instead of 84. With the 90 percent effective vaccine and Interaction Mode 1, the epidemic terminates at 235 days instead of 306.

## Discussion

We start from Figure 1, top panel. This demonstrates the desirable outcome with selective relaxation. Here, the disease is driven monotonically to elimination by allowing free interaction of vaccinees with each other while keeping tight controls on all other interactions. This elimination is not a “herd immunity” effect. The Figure shows that transmission levels are negligible post 250 days, at which time 1,50,000 people have been vaccinated, of whom 1,35,000 are successfully immunized (vaccine efficacy is 90). Add to this the 30,000 recovered cases and we have a total of 1,65,000 people who are immune when the transmission effectively halts. This amounts to 55 percent of the population, less than the 67 percent required to achieve “herd immunity” with a virus having an *R*_0_ of 3. Rather, our elimination drive is a vaccine-assisted variant of the auto-containment strategy which has worked successfully in countries like Australia, New Zealand and Taiwan. In countries with denser population, higher susceptibility etc, auto-containment by itself has proved impossible. However, vaccine-assisted elimination is much more feasible than containing the disease via NPI alone. The immediate lifting of restrictions for vaccinated people enables a steady expansion of social and economic activities during the elimination drive. Thus, selective relaxation with a highly effective vaccine features society heading towards normal and transmission towards zero – a win-win situation.

Figure 1, bottom panel shows a potential negative outcome of selective relaxation. Here, the vaccine efficacy is low so the high vaccinee—vaccinee interaction acts like a reopening measure. When sufficient susceptible vaccinees are available (at around *t*=350), the reproduction number exceeds unity and there is a wave of cases. We can see that the second wave occurs entirely in vaccinated middlers and seniors, and in minors who again get exposed by these people. From a practical viewpoint, the gradual rise in weekly cases around the 50^th^ week acts as a warning for an imminent second wave and signals the need for a stronger level of NPI. However, we would like to prevent such a situation beforehand; although the initial phases of the top and bottom panels of Figure 1 appear disturbingly similar, they have one significant difference. In the top panel, the bars corresponding to vaccinated cases (magenta, violet) are barely visible at any time during the initial decrease of cases while in the bottom panel, they are clearly present during this phase. A high prevalence of vaccinated cases during the initial deceleration of the pandemic can act as an early warning for a second wave and indicate the need for reimposing restrictions on vaccinee—vaccinee interactions.

In both these plots, the periodic five percent increments in the derate *δ* have a nuisance value. As mentioned in the Results Section, these increments slow down and can even destabilize the elimination drive. We have implemented this feature since it is very likely that as case counts decrease and vaccinees receive immediate clearance, non-vaccinees are also going to start bending the rules. If the data shows that these excursions are having a deleterious effect, then the public health authorities will have to intervene; otherwise the violations can be ignored.

What Figures 2-3 show is that for many vaccine-related parameters, there exists a sharply defined cutoff value above which the infection control performance is qualitatively similar to the ideal vaccine counterfactual and below which it is qualitatively similar to the no vaccination counterfactual (the death tolls though are much lower because the vaccine is highly effective against death even if not against case). The cutoff efficacy appears to be around 80 percent, a benchmark achieved by some of the currently available vaccine candidates but not by others. The cutoff vaccination rate in Figure 3 is about 400 doses/day, which is an easily achievable target. Once the cutoff is achieved, additional improvements in vaccination performance bring about only incremental effects in the overall disease management.

Finally, we have also found that varying *β* (the fraction of vaccines preferentially allotted to seniors) has a small effect on the overall case trajectories. In general, reducing *β* tends to reduce the duration and the cumulative caseload but increase the number of deaths. This is in line with the near-universal policy of vaccinating vulnerable people first, and it lends credence to the model predictions.

The **limitations** of the analysis come from the various assumptions in the model. One set of drawbacks is common to any lumped-parameter or compartmental model. This is that when the absolute number of cases becomes very low, the model ceases to remain valid – the deterministic evolution is replaced by a stochastic process. Hence, predictions regarding the end-stage of the outbreak, especially the time elapsed until elimination, might not be accurate. This apart, we have tried wherever possible to ensure that errors either cancel each other or occur on the side of caution. For example, the initial conditions feature a high case rate but no pre-immunized people; both of these militate against an elimination bid. We have taken minors to be as susceptible and transmissible as adults even though some evidence^32^ suggests that susceptibility and transmissibility might actually be lower for this group. We present a detailed discussion of the model assumptions and their effects in §2 of the Supplementary Data.

## Conclusion

In this Article, we have identified immediate and preferential relaxation of restrictions for vaccinees as a feasible path to the elimination of the terrible pandemic called COVID-19. This path features a continuous growth of economic and social activities during the vaccination drive. We hope that the incentive of immediate benefits will also induce people to get vaccinated and hence automatically combat vaccine hesitancy. With selective relaxation, and with current encouraging vaccine efficacies, we find a timeframe of eight to ten months before transmission reduces to negligible levels.

While our primary finding and its associated message are hopeful, there are also some cautionary takeaways. In particular, a 60-70 percent effective vaccine does **not** appear to be adequate for issuing immunity passports. Until and unless high-efficacy vaccines are widespread, research on improving vaccine efficacy should be pursued at maximum priority. “In the field” efficacy estimations should continue for all approved vaccines, especially to identify socio-demographic determinants of efficacy, if any.

In conclusion, the initiation of vaccination drives marks the beginning of the end of humanity’s struggle against COVID-19. Our immediate objective over the remaining few months of this battle has to be to minimize the caseloads, death tolls and socioeconomic disease burden. We hope that the prescription we have suggested here may prove effective in this respect.

## Data Availability

All data referred to in the Article is available publicly.

## Acknowledgements

We are grateful to the anonymous Reviewer for suggesting that we replace the original single-component model with an age-structured model, and use it to investigate the effects of minors’ ineligibility for vaccination. We also appreciate the Reviewer’s directing us to Mossong, Hens, Jit et. al.^23^ for the contact matrix.

## Conflict of interest statement

We have NO conflict of interest.

## Funding statement

We have NOT received any funding for this study.

## Author contribution statement

All of us have contributed equally to the manuscript.

## SUPPLEMENTARY DATA

In this Supplement we cover several issues which could not be treated in the Article proper due to lack of space. Throughout, a figure or table numbered “n” always refers to the Article proper while a figure or table numbered “Sn” refers to this document. The same holds for reference. Since labelled display equations exist in this Supplement alone, we have tagged them with numbers only and no “S” prefix.

### §1 MODEL DERIVATION AND EQUATIONS

We begin with a very brief recap of the model proposed in our prior work [21].

#### RECAPITULATION

Like every lumped-parameter or compartmental model, ours is applicable in any region with homogeneous mixing among its inhabitants, such as a neighbourhood, town, village, or smaller city. Metropolitan cities may need to be partitioned into several regions, depending on internal connectivity. The model treats the transmission of disease as a process of interaction between at large cases and susceptible targets; its underlying philosophy is

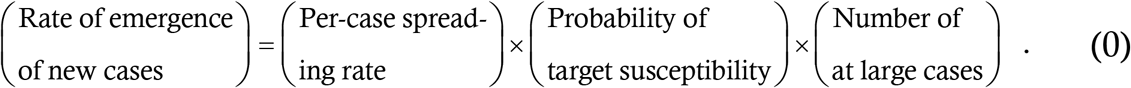

Defining *y* (*t*) as the cumulative case count, the left hand side above is d*y*/d*t*. The per-case spreading rate, which we call *m*_0_, is the product of two quantities – the rate *q*_0_ at which a random person (and hence an at large case who is unaware of infectious nature) interacts with other people, and the probability *P*_0_ that an interaction with a susceptible target results in a transmission. *q*_0_ is governed by the degree of social restrictions in place while *P*_0_ is determined by masking and sanitization; collectively, *m*_0_ embodies the effects of non-pharmaceutical interventions [21]. *q*_0_ happens to be available as a parameter from Literature; this fact will play an important role later. The target susceptibility probability factors in the immune response to the disease; with permanent immunity (and some plausible approximations), it takes the form 1 − *y*/*N* where *N* is the region’s total population. The number of at large cases has the following mathematical expression :

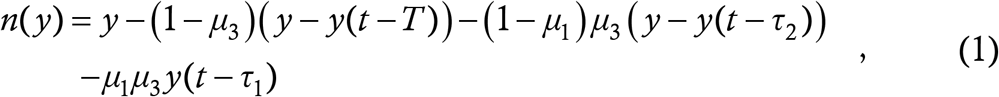

where *μ*_1_ is the fraction of cases who are asymptomatic, *μ*_3_ is the fraction of cases who escape from contact tracing, *T* is the time for which contact traced cases remain at large, *τ*_1_ is the time for which untraced asymptomatic cases remain transmissible and *τ*_2_ is the time for which untraced symptomatic cases remain transmissible and at large before manifesting symptoms and (at least so we assume) seeking quarantine.

Putting all this together, we arrive at the retarded logistic equation

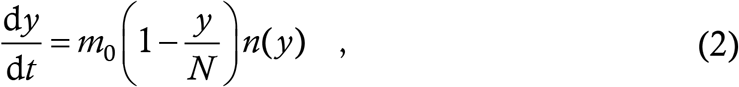

as the final form of the one-component epidemic model without vaccination. Equation (2)uses delays rather than inverse-rates to express infection durations, which enables it to make very realistic predictions. For further details of derivation, we must refer to our prior study [21].

#### ONE-COMPONENT VACCINATION MODEL

Here we add vaccination without age-structuring to the basic model (2); once this is done, the age-structured model will follow easily. We define three dependent variables : *y* (*t*) the cumulative count of corona cases among un-vaccinated people, *z* (*t*) the cumulative count of cases among vaccinated people and *v* (*t*) the total number of vaccinated people. We now use the logical structure (0) to formulate the evolution equations for the disease; for conceptual clarity, we permute the terms on the right hand side as follows :

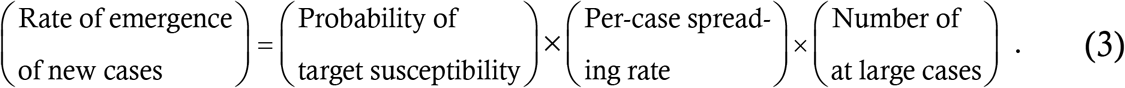

In this layout, the two terms featuring cases rather than targets are adjacent to each other.

We start from the interaction rates. In the most general case, there will be four interaction rates : *Q*^*a*^ the number of non-vaccinated people (targets) with whom one non-vaccinated person (and hence such an at large case) interacts each day, *Q*^*b*^ the number of non-vaccinated targets with whom one vaccinated case interacts each day, *Q*^*c*^ the number of vaccinated targets with whom a non-vaccinated case interacts each day and *Q*^*d*^ the number of vaccinated targets with whom a vaccinated case interacts each day. We have used capital *Q* rather than small *q* for a reason which will become apparent shortly, and have used superscripts for *a,b,c,d* rather than the more conventional subscripts for a reason which will emerge in the next Subsection. *Q*^*b*^ and *Q*^*c*^ are not necessarily equal; if a vaccinated person visits the houses of her ten unvaccinated friends who themselves remain confined at home then that contributes 10 to the vaccinee’s *Q*^*b*^ but only one to each of the non-vaccinees’ *Q*^*c*^.

Next, we argue that *Q* should be proportional to the total fraction of target population present i.e. *Q*^*a*^ and *Q*^*b*^ should be proportional to the total fraction of non-vaccinees while *Q*^*c*^ and *Q*^*d*^ should be proportional to the total fraction of vaccinees. To understand this, let us say Alfa is a vaccinee and she visits 10 friends per day. If the vaccine coverage is 20 percent then on average 8 of these friends will be unvaccinated and 2 vaccinated, so *Q*^*c*^ for Alfa will be 8 while *Q*^*d*^ for her will be 2; if on the other hand vaccination coverage is 80 percent then Alfa’s *Q*^*c*^ will be 2 and her *Q*^*d*^ will be 8. Since the population fractions of vaccinees and non-vaccinees change continuously, this dependence adds a time-varying component. We express this explicitly, writing

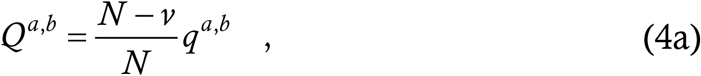

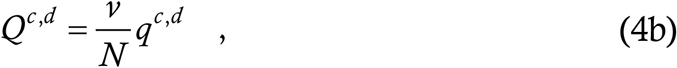

so that the *q*^*i*^’s become interaction parameters which can be obtained from Literature. Finally, we multiply each *q*^*i*^ by the transmission probability *P*_*i*_ to form the four spreading rates *m*^*a*^, *m*^*b*^, *m*^*c*^ and *m*^*d*^. The *P*_*i*_’s need not be the same for the four interaction types, since there might be heterogeneities in masking, handwashing etc.

To calculate the number of at large cases, we take the asymptomatic fraction *μ*_1_ = 4/5. We also take the contact traced fraction to be zero so that *μ*_3_ = 1. In Ref. [21] we have shown that contact tracing can capture only a small percentage of total cases if the asymptomatic fraction is high; moreover, contact tracing is managed by healthcare professionals many of whom are now re-deployed to vaccination drive. We use the parameter values [S1] *τ*_1_ = 7 and *τ*_2_ = 3, so that the function *n* gets defined as

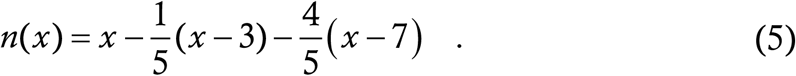

We assume that vaccinated cases have the same *μ*_1_, *τ*_2_ and *τ*_1_ as non-vaccinated ones (consequences of this come in §2), so that we can use this function *n* to count at large cases of both unvaccinated and vaccinated groups.

Now let us use (3) to formulate the equation for *y* (*t*), the unvaccinated cases. At any time, the total number of vaccinees is *v* and the total number of non-vaccinees is *N* − *v*. In this work, we assume that the disease as well as the vaccine confers permanent immunity (the disease with 100 percent probability and the vaccine only in those instances where it works), an assumption discussed in detail in §2. An unvaccinated person can be insusceptible only if s/he has already contracted and recovered from the disease; at any time the total number of recoveries (modulo the approximations of the previous Subsection) is *y*, the total number of non-vaccinees is *N* − *v* and so the probability that a random non-vaccinee is susceptible is (*N*−*v*−*y*)/(*N*−*v*), which is 1 − *y*/(*N*−*v*).

Cases arise in the unvaccinated group as a result of interactions whose targets are non-vaccinees, hence the relevant *m*’s in the *y* equation will be *m*^*a*^ and *m*^*b*^. We multiply *m*^*a*^ by the total number of at large unvaccinated cases and *m*^*b*^ by the total number of vaccinated cases, add the two and remember to account for the factor (*N*−*v*) / *N* from (4a) in the actual interaction rates which are *Q* rather than *q*. Putting all this together, we have

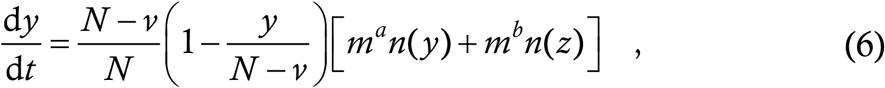

our first equation.

Similarly we can use (3) to formulate the equation for *z* (*t*). By the model assumptions, the vaccine confers sterilizing immunity with probability *η*, so at any time, the number of insusceptible vaccinees is *ηv* and the number of susceptible vaccinees is (1−*η*)*v*. Among the latter, *z* people have contracted and recovered from the infection so they are insusceptible as well. Hence, the total number of susceptible vaccinees is (1−*η*)*v* − *z* and the susceptibility probability is this divided by *v*, which is 1−*η* − *z*/*v*. The fraction (*N*−*v*)/*N* in (6) will now get replaced by *v*/*N*; moreover the terms *m*^*a*^ and *m*^*b*^ will get replaced by *m*^*c*^ and *m*^*d*^ since the target is a vaccinee. This yields

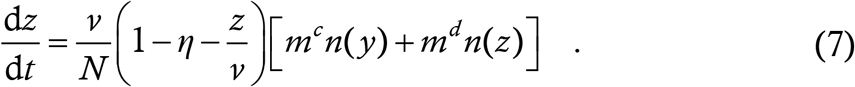

Finally, we need an equation for *v*. We assume that vaccination takes place at a constant rate *α* (people/day). The longest that the vaccination drive can continue is until all non-vaccinees have turned into either cases or vaccinees i.e. when *y* + *v* equals the total population. Thus, we have

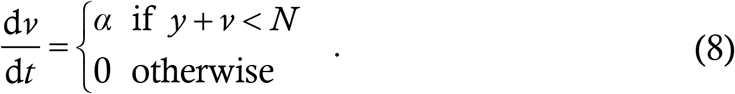

The stopping condition assumes that everyone is willing and able to receive the vaccine, which is sufficient for this single-population model.

Equations (6-8) constitute the one-component vaccination model; we now extend it to account for age-structuring.

#### AGE-STRUCTURED VACCINATION MODEL

As discussed in the Article proper, we consider three age groups (*a*) Group 1 aged 0-19 or “minors”, (*b*) Group 2 aged 20-64 or “middlers” and (*c*) Group 3 aged 65+ or “seniors”. Minors cannot be vaccinated so these cases account for a single dependent variable; in the other two categories we need three variables each corresponding to unvaccinated cases, vaccinated cases and vaccinees. Let the variable *y*_1_ denote minor cases, *y*_2_ unvaccinated middler cases, *z*_2_ vaccinated middler cases, *v*_2_ vaccinated middlers, *y*_3_ unvaccinated senior cases, *z*_3_ vaccinated senior cases and *v*_3_ vaccinated seniors. Let *N*_1_, *N*_2_ and *N*_3_ denote the total populations of minors, middlers and seniors respectively.

So far as parameters are concerned, the Literature yields nine interaction rates *q*_*ij*_ denoting the rate (per day) at which a person belonging to group *i* interacts with people belonging to group *j*. This can be converted to *m*_*ij*_ via multiplication by a relevant probability. Then, each *m*_*ij*_ can have a different value depending on whether the interacting cases and targets are vaccinated or otherwise. Once again, we use the superscripts *a,b,c,d* as in (6,7) to denote the different possibilities. *m*_11_ has only one value because all minors are unvaccinated; *m*_*i*1_ or *m*_1*i*_ can have two values depending on the vaccination status of the adult while *m*_*ij*_ for *i,j* = 2,3 can have four values each.

In addition to the vaccination rate *α*, we now need another parameter *β* (introduced in the Article proper) denoting the fraction of vaccines preferentially allotted to seniors. We assume that at the start of the vaccination drive, *βα* vaccines per day are distributed to seniors and (1−*β*)*α* vaccines per day to middlers. When any of the groups reaches saturation, all *α* vaccines are then diverted to the other group.

Given this, we can present the vaccination model. We have

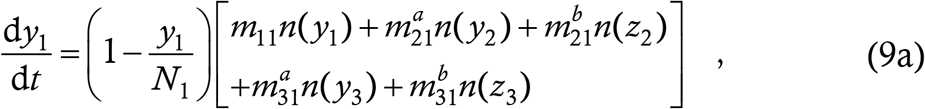

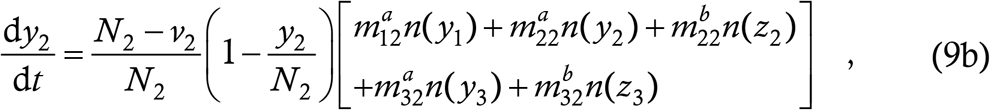

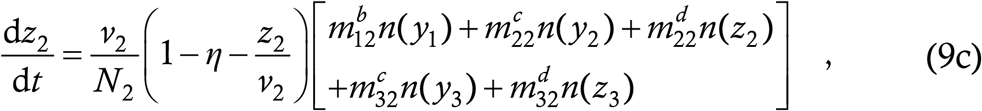

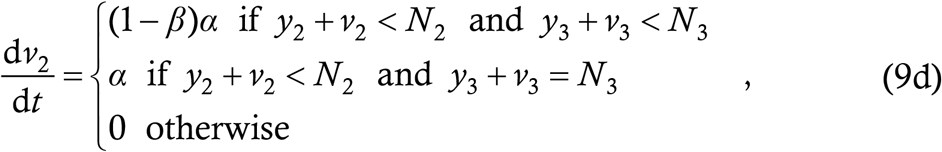

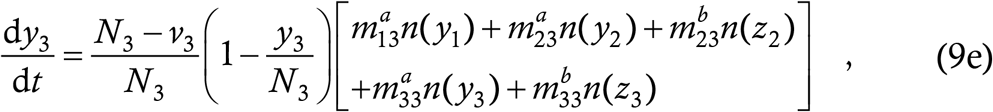

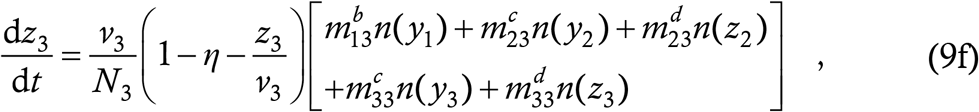

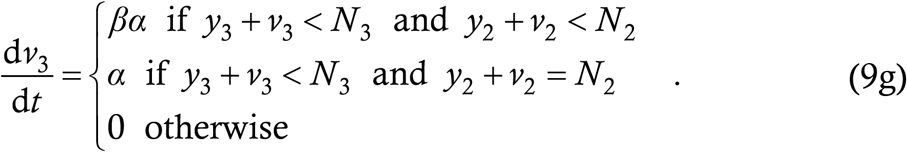

This is the equation set we simulate in the Article proper. We now describe the derivation of the various parameter values used in the simulation runs.

#### PARAMETER DERIVATION

The first raw material we need for this is a population dataset. As a representative population, we use recent data from USA [22], which has a total population of 328·2 millions. In Table S1, we show the total number of people in various age brackets, together with some percentage compositions.

**Table S1 :** Demographics of the USA population [22]. The first three columns are self-explanatory. The fourth column shows the population of the concerned age bracket as a percentage of the total population of the group, for example age 0-4 accounts for 23·99 percent of the total population of Group 1. The fifth column shows the population of the group as a percentage of that of the whole country.

| Group | Age | Population<br>(millions) | Age : Group<br>(Percent) | Group : Total<br>(Percent) |
| --- | --- | --- | --- | --- |
| 1 | 0-4 | 19.58 | 23.99 | 24.87 |
|  | 5-9 | 20.19 | 24.73 |  |
|  | 9-14 | 20.80 | 25.48 |  |
|  | 15-19 | 21.06 | 25.80 |  |
| 2 | 20-24 | 21.63 | 11.23 | 58.66 |
|  | 25-29 | 23.50 | 12.21 |  |
|  | 30-34 | 22.43 | 11.65 |  |
|  | 35-39 | 21.73 | 11.29 |  |
|  | 40-44 | 19.92 | 10.35 |  |
|  | 45-49 | 20.40 | 10.60 |  |
|  | 50-54 | 20.48 | 10.64 |  |
|  | 55-59 | 21.87 | 11.36 |  |
|  | 60-64 | 20.57 | 10.68 |  |
| 3 | 65-69 | 17.46 | 32.29 | 16.47 |
|  | 70-74 | 14.03 | 25.95 |  |
|  | 75-79 | 9.65 | 17.85 |  |
|  | 80-84 | 6.32 | 11.69 |  |
|  | 85+ | 6.61 | 12.22 |  |

Extrapolation from this dataset to the Notional City of population 3,00,000 has yielded *N*_1_ = 74,610, *N*_2_ = 1,75,980 and *N*_3_ = 49,410.

The next piece of raw material required is the contact matrix. For this we have used data from Ref. [23]. The authors of this study have conducted a survey to calculate the average number of people belonging to one age group with whom one person belonging to another age group interacts each day. This is of course *q*_*ij*_ except that Ref. [23] uses the 5-year age bands from Table S1 to generate a 15×15 contact matrix (it treats 70+ as the last bracket), while we need a 3×3 matrix. Ref. [23] reports on surveys conducted in different countries; we use the UK as an example since the mean interaction rate there is very close to the mean over all the considered countries. Specifically, we use the data from Table S8.4a in the Supporting Information of Ref. [23].

We demonstrate the reduction of contact matrix from 15 square to 3 square using one example – the contact rate *q*_11_. First, we reproduce a part of Table S8.4a of Ref. [23] as Table S2 below.

**Table S2 :** An extract from Table S8.4a of Ref. [23]. It shows the average number of daily contacts (“targets”) belonging from different age groups along the columns, as reported by each person (“case”) belonging from different age groups along the rows.

| **Target\Case** | **0-4** | **5-9** | **10-14** | **15-19** |
| --- | --- | --- | --- | --- |
| **0-4** | 1.92 | 0.65 | 0.41 | 0.24 |
| **5-9** | 0.95 | 6.64 | 1.09 | 0.73 |
| **10-14** | 0.48 | 1.31 | 6.85 | 1.52 |
| **15-19** | 0.33 | 0.34 | 1.03 | 6.71 |

As per Table S2, a case aged 0-4 interacts every day with 1·92 targets aged 0-4, 0·95 targets aged 5-9, 0·48 targets aged 10-14 and 0·33 targets aged 15-19. All these are Group 1 targets, so we can simply add along the column to find that a case aged 0-4 interacts with 3·68 Group 1 targets every day. Similarly, adding along the other columns yields that a case aged 5-9 interacts with 8·94 Group 1 targets, a case aged 10-14 interacts with 9·38 Group 1 targets and a case aged 15-19 interacts with 9·20 Group 1 targets every day. Now, to obtain the Group 1—Group 1 interaction rate, we must weight these numbers by the population fractions from column 4 of Table S1 and add them up. This yields *q*_11_ = 7·85. This tedious operation repeated over all groups yields the contact matrix which we have displayed in the Article proper and reproduce here for clarity :

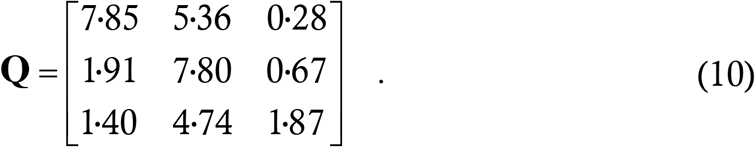

Again, this matrix is not expected to be symmetric – consider a 25-year old nurse visiting ten 80+ year-olds in an assisted living facility. We assume that contact reductions (for example lockdown) take place in such a manner as to reduce all elements of **Q** by the same factor.

The next parameter to account for is the raw transmission probability *P*_0_, in the absence of masking or other interventions. This is difficult to measure a priori so we shall obtain it indirectly, by tying it up to the reproduction number *R*_0_ of the disease, which is a measurable quantity [25–28]. The direct calculation of *R*_0_ from the system (9) is a hair-raising task, but we can get the value from a qualitative argument.

*R*_0_ is defined as the number of people whom one case infects on average when everyone is susceptible and there are no external interventions. The first row of **Q** yields that a Group 1 case interacts with 7·85+5·36+0·28 = 13·49 targets every day; he transmits the disease to 13·49*P*_0_ of them. Similarly, a Group 2 case transmits the disease to 10·38*P*_0_ people every day and a Group 3 case transmits to 8·01*P*_0_ people every day. At the start of the outbreak, we assume that cases occur in the three groups in proportion to their populations; weighting the groups’ contributions by the percentages in the fifth column of Table S1 and adding yields that one case transmits to 10·76*P*_0_ targets per day on average. By the model assumptions, 80 percent of all cases are asymptomatic and transmit for 7 days while 20 percent are symptomatic and transmit for 3 days, so that an at large case transmits for 31/5 days on average. Thus, a case transmits the disease on average to (10·76*P*_0_) × (31/5) people which is 66·71*P*_0_ people. Hence we claim *R*_0_ = 66·71*P*_0_.

Having obtained this expression by crook rather than by hook, we must check its validity by numerical simulation of (9). If the condition is correct, then the epidemic ought to take off if *P*_0_ > 1/66·71 and die down otherwise. Simulating with different initial conditions, we find that the transition occurs at a *P*_0_ value of about 96-99 percent of the calculated one. Hence our expression for *R*_0_ is correct and we can use it to define *P*_0_ as

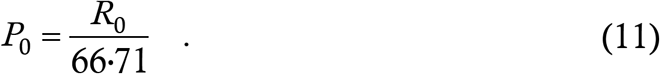

For normal life, *R*_0_ can be 2, 3 or 5 depending on the viral strain, and the corresponding *P*_0_ is obtained through (11). We define

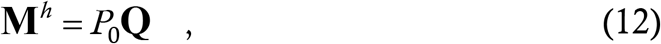

as the matrix of the spreading rate *m*_*ij*_’s during normal life. During pandemic life, each *m*_*ij*_ can have upto four values as we can see from (9); to simplify the parameter space we define the matrix

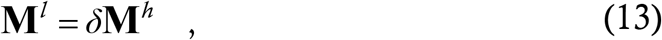

where the derate *δ* lies between 0 and 1, and stipulate that 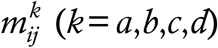 can be either 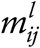 or 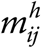 and nothing else. The deration takes care of contact reduction as well as transmission probability reduction through measures such as masking. Thus, for low-transmissibility strain we use the values *P*_0_ = 2/66·71 and the initial derate *δ*_0_ = 0·489, for consensus strain we use *P*_0_ = 3/66·71 and *δ*_0_ = 0·327 and for rogue strain we use *P*_0_ = 5/66·71 and *δ*_0_ = 0·196. These values of *δ*_0_ were chosen so as to keep the case rate curve closest to horizontal during the initial week or so of the vaccination drive.

Finally, we explain how we implement the two interaction modes in the analysis. For Mode 1, we choose

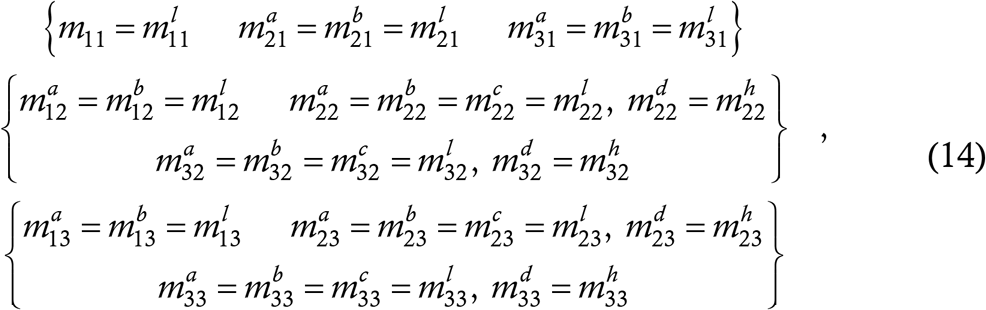

while for Mode 2 we choose

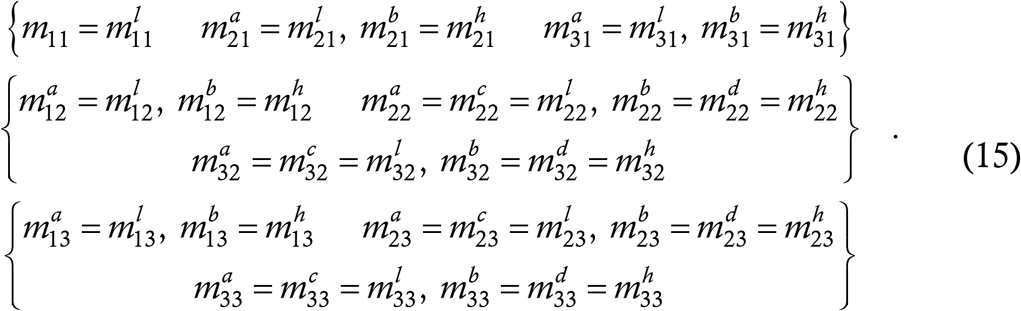

Equations (14,15) look much more cumbersome than the concepts which they embody.

For the groupwise mortality rate, we use data from Ref. [31]. Table S3 in the Supplementary Information of this paper gives the mortality rates for the age brackets considered in Table S1 here; we weight them by the percentages in the fourth column of this table to obtain the mortality rates used in the text.

Our final concern is with the initial and terminal conditions. We have chosen the initial conditions to generate a steady 300 cases/day for the first seven days, with cases divided among the three groups approximately in proportion to their population; this amounts to a daily case rate of 1/1000^th^ the total population and is very high. For termination condition, we define the active case count at time *t* to be

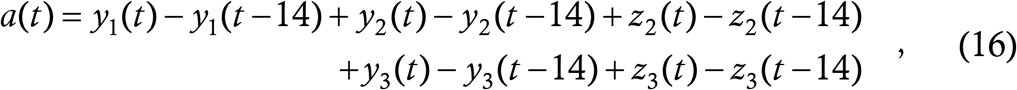

and stop the run if *a*(*t*) < 1 for 14 consecutive days.

### §2 MODEL ASSUMPTIONS AND THEIR EFFECTS

Here we discuss some of the assumptions and approximations inherent in the model and the effects which they have on the results. For assumptions built into in the baseline model (2), we cite Ref. [21]; here we consider only the approximations involved in extending (2) to form (9).

#### Vaccine immunity

As mentioned in §1, we have assumed that vaccine confers sterilizing immunity – i.e. complete immunity against contraction, symptoms and transmission of COVID-19 – with probability *η* and zero immunity against contraction and transmission with probability 1−*η*. Currently, the Pfizer and Moderna trials [1,2] have focussed only on reduction of symptomatic infections although, as we have mentioned in the Article proper but see no harm in repeating, a recent study from Israel [24] has found that Pfizer is highly effective against asymptomatic cases as well. The Oxford vaccine [3] has included asymptomatic cases in the phase 3 analysis and found only a 30 percent reduction in asymptomatic infections in the vaccine group relative to the placebo group (compared to a 60 percent reduction in symptomatic infections). However, the absolute number of asymptomatic cases detected in this study is quite small and more data needs to be collected on this issue.

If the Oxford results are indicative of a general trend, then our assumption of equal efficacy against asymptomatic and symptomatic infection will result in an undercounting of cases since our model will yield fewer asymptomatic vaccinated cases than reality. This is counterbalanced by our assumption that vaccinated cases have the same transmission properties (transmissibility and duration) as unvaccinated ones. It is possible (and intuitive) that vaccine cases will actually have lower viral loads and faster recovery period, which will cause reality to undershoot the model prediction. Indeed, a second study from Israel [S2] reports positive news on this front. It finds that cases who have contracted the disease 12-28 days after receiving the first dose of Pfizer vaccine feature a fourfold reduction in viral load compared to unvaccinated cases. To the best of our knowledge, so far, there is nowhere near enough information which can enable us to accurately determine more appropriate parameter choices. However, when such information does become available, it will not be difficult to incorporate it into the model by changing the values of *η* and of *μ*_1_, *τ*_1_, *τ*_2_ and the *m*’s for vaccinated cases.

While calculating the death tolls, we have assumed that vaccination reduces the mortality rates by a factor of ten. Currently, a value for this factor is unknown; all that is known is that in the phase 3 trials of any of the vaccines developed so far, there have been zero COVID-19 deaths in the vaccine groups.

A further assumption we have made is that the immunity conferred by the disease as well as the vaccine is permanent. This assumption is valid so long as the immunity duration is longer than the evolution time of the outbreak, which is less than a year in the unimodal solutions that we have obtained. For the disease itself, antibodies as well as cellular immune responses do seem to be durable over at least a 6-7 month period, the longest studied so far (a mini-review of literature on this topic appears in Ref. [S3], while Ref. [S4] is a recent update). As for the vaccine, Moderna [S5] and ICMR/BB [5] have reported durable immune responses for at least 3 months, with the titre profiles being similar to those generated by symptomatic COVID-19 infection. Time alone will tell us the durability of vaccine immune response, but so far we see no reason to deviate from the permanent immunity assumption. Note however that the bimodal solutions obtained with selective relaxation last indefinitely, and how the immunity business will play out in that scenario is anybody’s guess. We hope that in reality we shall not have to see the ugly face of the bimodal solution.

#### Initial and terminal conditions

The assumption that there are zero pre-existing cases at the start of the vaccination drive is an underestimate; in some regions at least, a significant fraction of the population has already been immunized. In other regions however, the immunized fraction might not be too large. Pre-existing recoveries can influence the case trajectories in two ways : (*a*) for given interaction parameters, it can make the actual reproduction number lower than the model and hence terminate the epidemic faster and with lower caseload, (*b*) it can achieve the reproduction numbers of our simulations at higher levels of mobility and hence equal our infection control performance at a lower level of intervention. The high initial case rate will tend to generate a large number of vaccine cases at the start and push up the vaccine fault ratio. Thus, both the starting case count and case rate are chosen to generate a maximally unfavourable scenario. The assumption of 100 pre-existing middler and senior vaccinees has no impact other than to prevent division by zero when calculating the fault ratio.

The terminal condition of less than one active case for a sufficiently long time is an eminently plausible measure of the true end of the outbreak. The number 14 (twice) in the definition of the condition might appear somewhat arbitrary. The choice is harmless since changing that number changes the cumulative case counts by minuscule amounts. At any rate, when the absolute number of cases is very low, a lumped-parameter model breaks down. All that one can talk about are probabilities, and for that one needs an agent-based model. Our model (and **any** other differential equation model) is good only for predicting when transmission will have become significantly reduced, and for that any physically plausible termination condition is adequate. We expect that the stochastic tail-phase of the outbreak will not add too many cases to our calculated totals; however it might prolong the epidemic significantly.

#### Vaccination fault ratio

The question we want to address is “If I receive the vaccine and party with other vaccinees, what is the probability that I shall actually contract the disease during the evolution of the outbreak ?” The guess answer 1−*η* is a gross overestimate. To see this, consider an individual vaccinee, whom we call Bravo. Vaccine efficacy of 90 percent implies 10 percent failure probability which does not sound very small. However, 10 percent is the probability that Bravo catches the disease **given** an exposure. Transmission is a two-person process – if Bravo interacts only with other vaccinees, then the probability that they have the disease and can expose Bravo to the pathogen also reduces to (approximately) 10 percent. Bravo’s total contraction probability therefore reduces to approximately 1 percent i.e. the disease contraction probability is quadratic and not linear in the vaccine failure probability. This argument is slightly hand-waving in character but it does convince us, independent of the maths, that the contraction probability will be much lower than the complement of the efficacy.

There is no single metric in fact which can help us to answer the above question. An approximate indicator will be the ratio of the total number of vaccinated cases to the total number of vaccinees. However, this index will be artificially lowered by the fact that during the tail-phase of the epidemic, there are hardly any new cases but lots of new vaccinees. Hence we have opted to evaluate the ratio at every point during the disease evolution and report its maximum value as the vaccination fault. It is comforting that for the unimodal solutions, the fault evaluates to less than 1 percent.

#### Details of structuring

Here we used a social structure based on contact rates in UK superposed on demographics of USA. The question arises as to what extent the results are dependent on the choice of structuring. Moreover, instead of three age groups, suppose we had used say five or fifteen, then would the results have been different ?

To answer this question, we consider the unstructured model (6-8) and compare its predictions with those of (9) for the same parameter values. For the first scenario, we take Mode 1, consensus strain. To achieve this, we use *m*^*a*^ = *m*^*b*^ = *m*^*c*^ = *m*^*l*^ = 0·163 and *m*^*d*^ = *m*^*h*^ = 0·489 in (6-8) [a simple calculation along the lines of that in §1 here provides these values]. We again increment *m*^*l*^ in steps of 5 percent every 50 days. With 90 percent vaccine efficacy, we find that the epidemic ends in 205 days with 16,000 cases and a vaccination fault ratio of 0·18 percent. The time trace of evolution is shown in Figure S1 below.

**Figure S1 :**
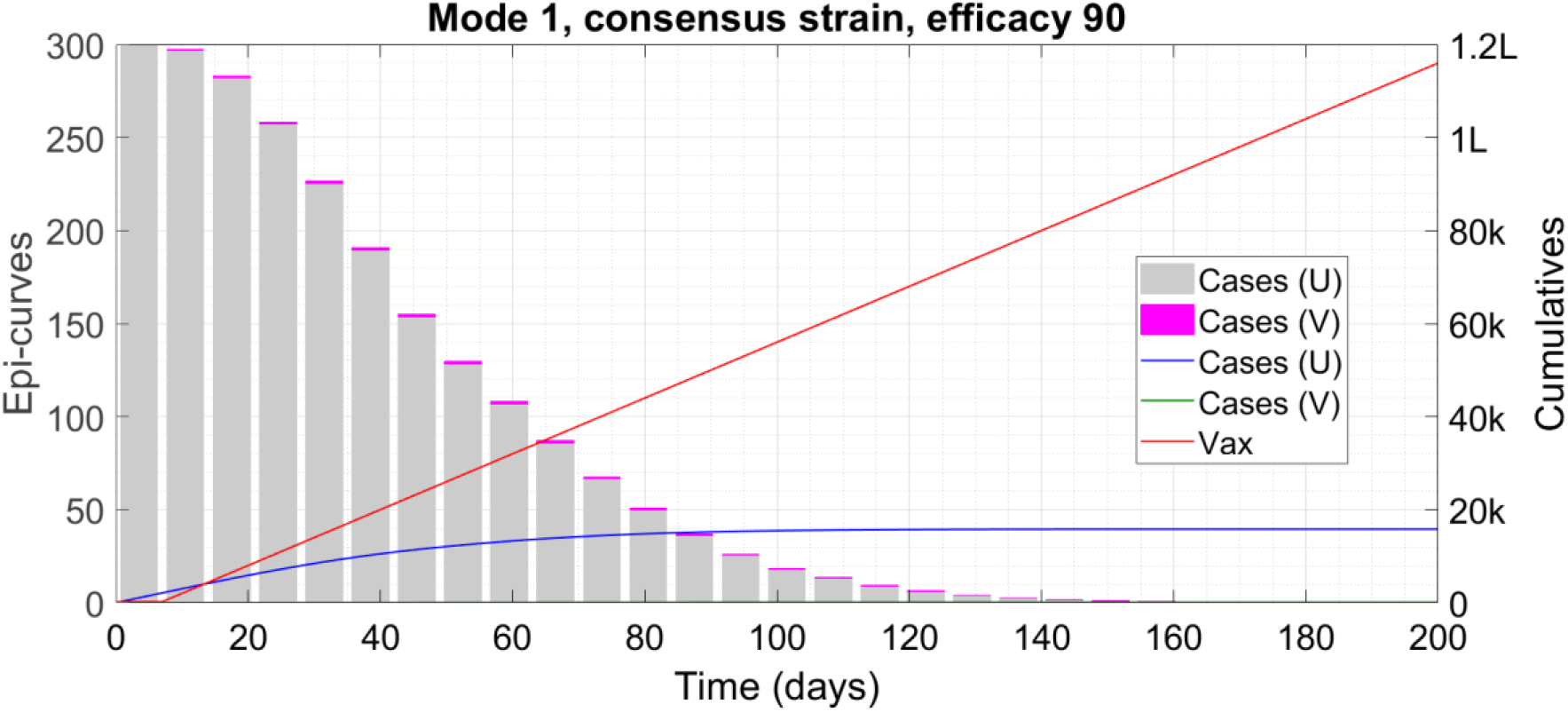
Time trace of epidemic evolution showing unimodal solution. We display cumulative counters such as caseload and vaccinee count as lines associated with the right-hand y-axis and the weekly case counts or epidemiological curve (epi-curve) as bars associated with the left-hand y-axis. We have scaled down the weekly counts by a factor of 7 so that the envelope of the bars coincides with the time derivative of the cumulative cases. The symbol ‘k’ denotes thousand and ‘L’ hundred thousand.

With 60 percent efficacy on the other hand, the outbreak ends at 908 days with total 44,000 cases and a vaccination fault ratio of 8·9 percent. The evolution is shown in Figure S2 below.

**Figure S2:**
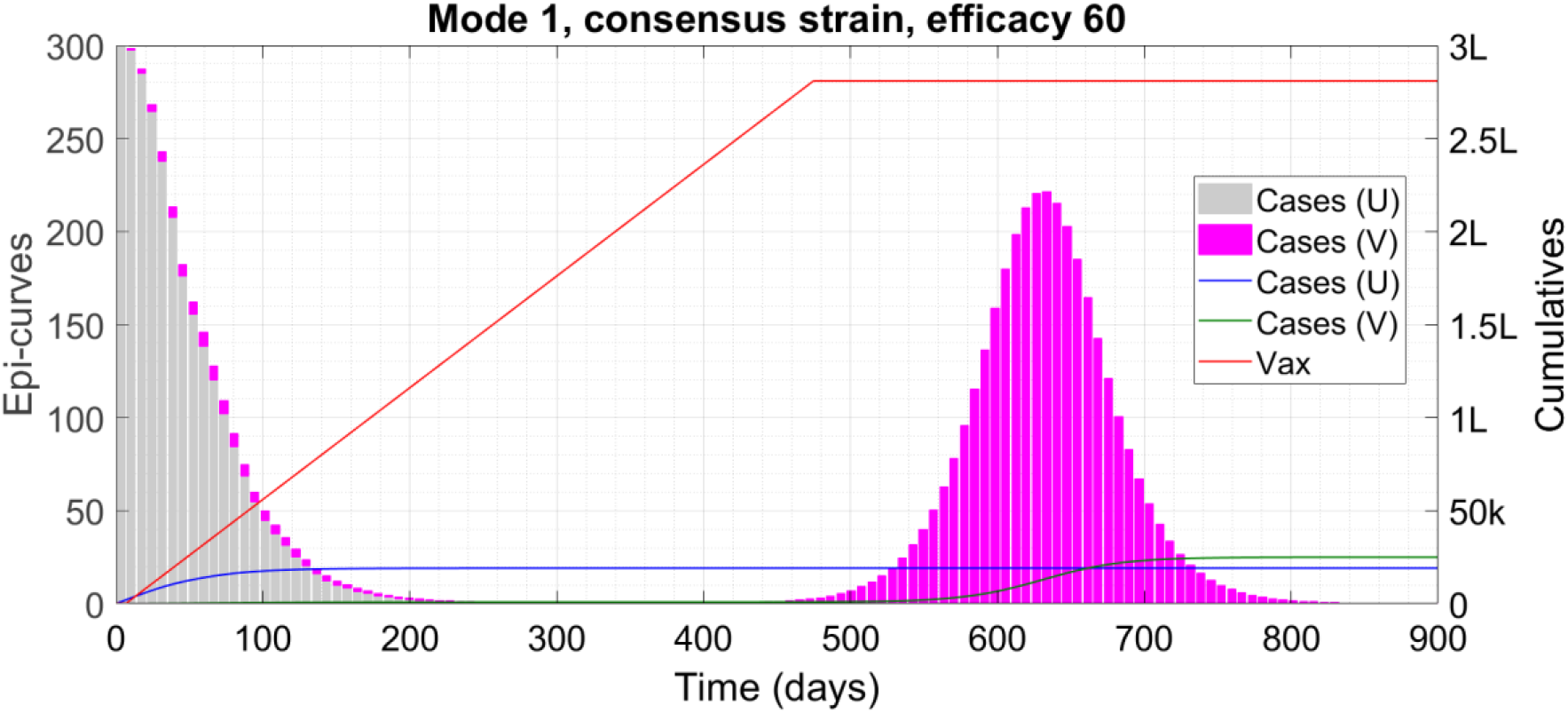
Time trace of epidemic evolution showing bimodal solution. We display cumulative counters such as caseload and vaccinee count as lines associated with the right-hand y-axis and the weekly case counts or epidemiological curve (epi-curve) as bars associated with the left-hand y-axis. We have scaled down the weekly counts by a factor of 7 so that the envelope of the bars coincides with the time derivative of the cumulative cases. The symbol ‘k’ denotes thousand and ‘L’ hundred thousand.

We see that the general trends of unimodal elimination and bimodal long-run epidemic remain the same. In the unstructured model, the overall durations and caseloads are less than in the structured model, which is very plausible since the latter model includes a class (minors) who have high interaction rates but are ineligible for the vaccine.

As a final comparison, we repeat the plot of Figure 2, top panel (caseload and duration vs efficacy for Interaction Mode 1 with the rogue strain) using the unstructured model instead of the structured one. We show the result in Figure S3.

**Figure S3 :**
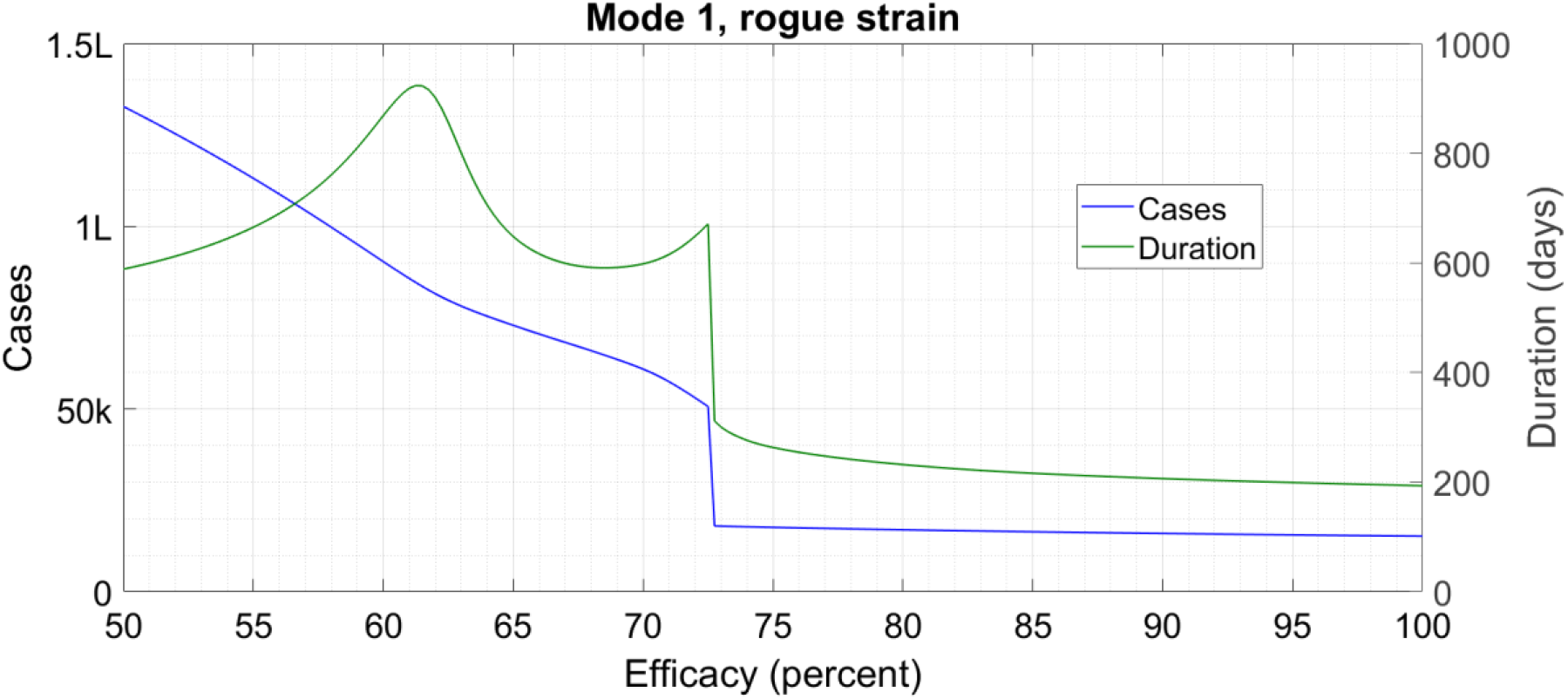
Caseload (left-hand y-axis) and duration (right-hand y-axis) as a function of vaccine efficacy for one combination of interaction mode and viral strain. The jump in the curves corresponds to a transition from a unimodal to a bimodal solution profile. The symbol ‘k’ denotes thousand and ‘L’ hundred thousand.

Once again, there is a sharp change in the curve when the unimodal solution changes to the bimodal one; the cutoff point reduces from 78 percent in Figure 2 to 73 percent in Figure S3, consistent with the simplifications in the model. As we have mentioned in the Article proper, we have taken minors to be as susceptible and transmissible as adults, ignoring evidence [32] which suggests that this might not be true. If minors do have lower susceptibility and transmissibility, then the actual case counts will be lower than the model predictions.

Overall, the comparison between the unstructured and structured models shows that the predictions remain generally similar in both situations, with the differences lying in the details. This convinces us that further refinement in the structuring or small changes in parameter values are not going to have a significant impact. During future refinement of the model, one further feature which can be taken into account is interaction-structuring. This factors in that some people such as shopkeepers, bankers and bus conductors are forced to interact with dozens every day for professional reasons while others like researchers who work on modeling studies lead much more reclusive lives (especially in the present situation). Preferentially vaccinating people of the former category, a strategy considered in Refs. [10–13], can lead to significant gains in duration and caseload.

#### Vaccine hesitancy

This is a phenomenon which our model does not account for explicitly. However, in most of the successful runs in Table 1, we can see that transmission is significantly reducing within about 250 days, when about 1,50,000 vaccines have been administered. This amounts to approximately 2/3 of the eligible population, so it automatically allows scope for a considerable amount of hesitancy. Even if hesitancy causes the vaccination drive to stop when transmission is reduced but not fully eliminated, the epidemic will continue on a downward spiral so long as the restrictions on non-vaccinees are kept in place. Moreover, the disparity in social lives of the vaccinees and non-vaccinees should cause many of the hesitant people to capitulate. Hence, vaccine hesitancy should not be too great a factor affecting the vaccine-assisted elimination of the disease.

### §3 SOCIOECONOMIC AND POLICY ASPECTS

With an effective vaccine, selective relaxation appears to be a quick and surefire path to elimination of COVID-19 in time while achieving maximum socioeconomic recovery. This process however may cause negative emotions between people who get vaccinated earlier and those who get vaccinated later. To the largest extent, there is nothing to be done about this rift – while vaccine allocation policies can come up with a priority order which maximizes the common good, it will be inevitable that one healthy young well-paid software engineer will get the shots and hence a ticket to freedom two months before another healthy young well-paid software engineer.

The negativity arising from such heterogeneity can be alleviated through a public information campaign – the impact of the virus itself is very heterogeneous and that is outside our control, so a bit of manmade inequality during the endgame phase is also tolerable. This is especially true since continuing blanket restrictions until the disease has been eliminated will entail tremendous economic losses. Nonetheless, to avoid unhealthy competition, employers and universities who arrange for vaccination of employees and/or students might wait to initiate the general vaccination drive (excluding frontline, high-interaction and high-risk people) until they have secured all the requisite doses.

During selective relaxation, public health authorities will have to ensure that minimal close and unmasked interaction occurs between vaccinees and non-vaccinees. To facilitate this, vaccinees may be given apparel or badges which prominently advertise their status. In situations where physical segregation of the two classes is impossible, like shops and restaurants, mask and separation requirements will likely need to remain in place for the vaccinees as well. If a chain store or eatery has multiple similar outlets in the same city, like McDonald’s, it might designate some as vaccinee only with no restrictions and others as common spaces with restrictions. It goes without saying that vaccinee-only entertainment venues have to immunize their staff before admitting customers.

We are aware that selective relaxation is harder to implement than collective relaxation or restriction. This is probably one of the reasons why all the modeling studies referred to in the Article proper consider only collective interventions. However, with a high efficacy vaccine, the socioeconomic gains from selective relaxation are immeasurable while the epidemiological gains from collective restriction are trivial. For example, consider a situation with 90 percent effective vaccine and consensus viral strain. With Interaction Mode 1, we have already seen that the epidemic ends at 306 days and 30,193 cases. If we instead employ a collective interaction mode where all spreading rates are 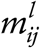 with no 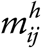, then the epidemic runs for 305 days accruing 30,161 cases. Thus, keeping an average of 90,000 people locked down during the vaccination drive would in this case be a “Useless Precaution”.

A plethora of results supports the assertion that a vaccine with 80 percent or higher efficacy can act as the basis for an immunity passport while a vaccine with efficacy in the 60s cannot, unless the virus transmission rate is already low. Hence, research into development of more efficacious vaccines should continue even as the early candidates are administered. The efficacies of existing vaccines should also be monitored for socio-demographic determinants if any. For example, if Vaccine A has 90 percent efficacy among middlers and 40 percent among seniors while Vaccine B has 70 percent across all age groups, then middlers should be given Vaccine A and seniors Vaccine B.

In conclusion, we look forward to the day when we can get an effective vaccine, doff our mask and return to life as we knew it, and we hope that this happy day arrives sooner rather than later.

## Notes

### Competing Interest Statement

The authors have declared no competing interest.

### Funding Statement

We have received NO funding for this study from any agency.

### Author Declarations

No IRB approval required for mathematical modeling study

